# Profiling cases with non-respiratory symptoms and asymptomatic SARS-CoV-2 infections in Mexico City

**DOI:** 10.1101/2020.07.02.20145516

**Authors:** Omar Yaxmehen Bello-Chavolla, Neftali Eduardo Antonio-Villa, Arsenio Vargas-Vázquez, Carlos A. Fermín-Martínez, Alejandro Márquez-Salinas, Jessica Paola Bahena-López

## Abstract

We profiled cases with non-respiratory symptoms (NRS) and asymptomatic SARS-CoV-2 assessed within Mexico City’s Epidemiological Surveillance System. We show that initially asymptomatic or NRS cases have decreased risk of adverse COVID-19 outcomes compared to cases with respiratory symptoms. Comorbidity and age influence likelihood of developing symptoms in initially asymptomatic cases.

## INTRODUCTION

Asymptomatic infections are a potential source of rapid SARS-CoV-2 spread^1^. Prevalence of pre-symptomatic/asymptomatic cases varies widely but represent a larger proportion of cases than previously anticipated ^2^. Controversies surrounding asymptomatic SARS-CoV-2 infections have focused on the distinction between pre-symptomatic and asymptomatic infection and its definitions; nevertheless, longitudinal data shows that only a fraction of initially asymptomatic infections develop subsequent symptoms ^3,4^. In Mexico, SARS-CoV-2 testing focuses on sampling high-risk cases with suspected COVID-19 by the presence of RS, which has allowed characterization of increased susceptibility for severe COVID-19 attributable to cardio-metabolic comorbidities ^5^; however, no data has been reported on cases with non-respiratory symptomatology (NRS) and asymptomatic SARS-CoV-2 infections, which are underrepresented under the current testing framework in Mexico. Lack of information regarding cases with NRS, or initially asymptomatic cases in Mexico is concerning, particularly since many studies suggest that contact tracing of asymptomatic infections will be pivotal to contain further SARS-CoV-2 spread and the likelihood of underreporting cases without respiratory symptoms but with NRS^6^. Here, we profile patients with NRS and initially asymptomatic SARS-CoV-2 infections to assess outcomes after follow-up using cases detected in Mexico City.

## METHODS

We analyzed data from the National Epidemiological Surveillance System in the Greater Mexico City area, an open-source dataset comprising daily updated cases tested using realtime RT-PCR to confirm SARS-CoV-2 using nasopharyngeal swabs according to the Berlin Protocol ^7^. For analysis, we propose two main symptom groups, which we confirmed using correspondence analyses to cluster symptoms with comparable similarity (**Supplementary Material**): RS included dyspnea, polypnea or cyanosis at initial evaluation, but we also included fever and cough in this group based on WHO criteria for suspected COVID-19 cases; whilst NRS included headache, myalgias, arthralgias, general malaise, abdominal pain, chest pain, conjunctivitis, irritability or vomiting. We defined cases with RS as those with ≥1 RS and any or none NRS. Cases with NRS had ≥1 NRS without any RS, whilst initially asymptomatic cases had no symptoms at the point of initial assessment. NRS and initially asymptomatic cases were assessed outside the established framework for suspected COVID-19 cases in Mexico by contact tracing of real-time RT-PCR confirmed SARS-CoV-2 infection and convenience sampling of cases without symptoms who requested an RT-PCR test for SARS-CoV-2; complete description of the Mexican testing framework is described in **Supplementary Material**. We also estimated 7-day rolling average positivity rates (PR) for RS, NRS and initially asymptomatic cases. Follow-up was conducted for all non-hospitalized cases up to 14 days; outcomes for all patients included hospital discharge due to clinical improvement or voluntary, ambulatory/treatment follow-up or death; additional information regarding follow-up is presented in **Supplementary Material**. Severe COVID-19 was defined as a composite of death, ICU admission or intubation. Complete description of statistical analyses and available variables are detailed in **Supplementary Material**.

## RESULTS

### Study population

As of 19^th^ August 2020, a total of 296,157 subjects were tested for SARS-CoV-2 in Mexico City. From 108,080 cases with confirmed SARS-CoV-2 infection (PR 36.5%), 14,657cases did not have RS (13.6%). Overall, 37,356 NRS cases had been evaluated, amongst whom 8,545 had confirmed SARS-CoV-2 infection (Overall PR: 22.9%); similarly, 35,175 initially asymptomatic cases were evaluated and 6,112 cases had confirmed SARS-CoV-2 infection (PR: 17.4%). Seven-day PRs had been steadily decreasing for all patient categories during the last few weeks (**Supplementary Material)**. NRS cases were younger, predominantly female and had lower prevalence of NRS compared to cases with RS. Initially asymptomatic cases were younger compared to RS cases, with similar trends for lower rates of comorbidities compared to both RS and NRS cases (**Figure 1**).

**Figure 1.**
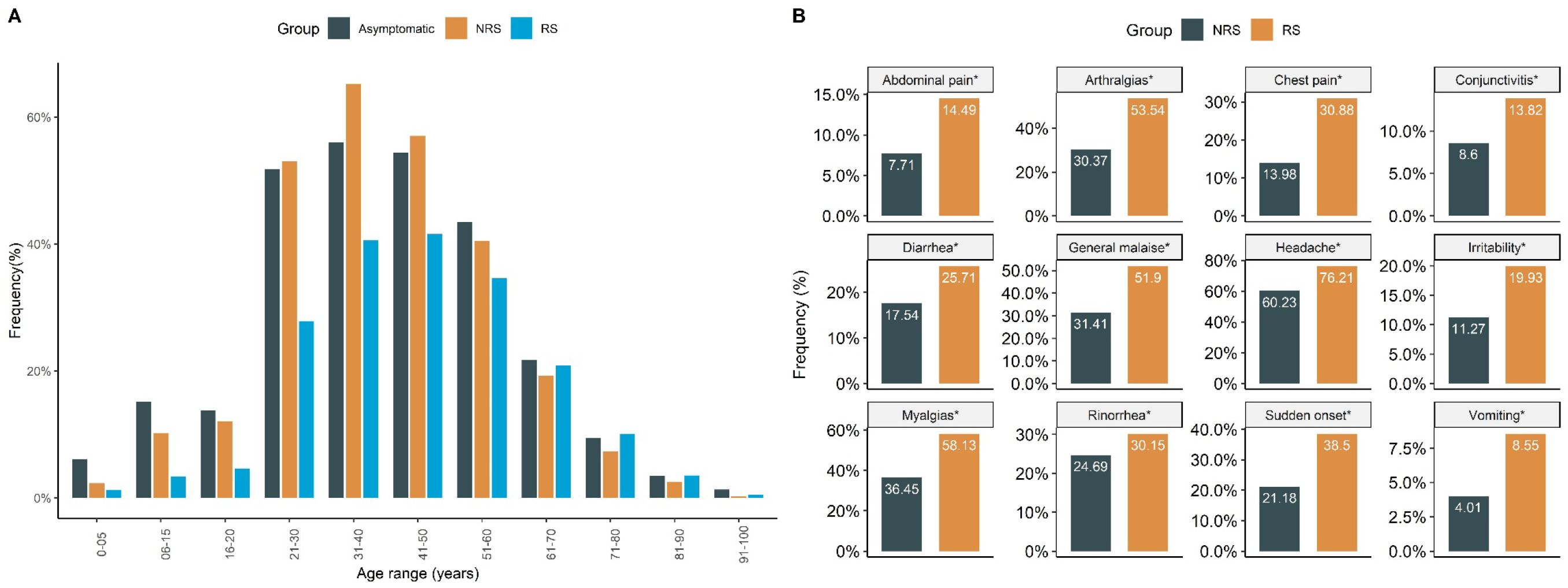
Histogram comparing case distribution according to age groups comparing cases with RS, NRS and asymptomatic SARS-CoV-2 infection, demonstrating the role of age in determining asymptomatic SARS-CoV-2 infections (A). We also show a comparison of the distribution of NRS related to SARS-CoV-2 infections in subjects with RS and NRS only. *p<0.001 *<u>Abbreviations</u>*: RS: Respiratory symptoms; NRS: Non-respiratory Symptoms; Sudden onset: Patient report of sudden onset of symptomatology.

### Profiling NRS cases with SARS-CoV-2 infection

Amongst NRS cases (n=8,545), 365 subjects were hospitalized (4.3%), 32 required mechanical ventilatory support (MVS) (0.4%), 32 required intensive care unit (ICU) admission (0.4%). Overall, 112 patients died (1.3%), of whom 24 deaths were ambulatory (0.3%) and 88 hospitalized (1.0%). Predictors that increased risk for severe COVID-19 in NRS cases included age >60 years, chronic obstructive pulmonary disease (COPD), diabetes, and chronic kidney disease (CKD); decreased risk of severe COVID-19 was associated with rhinorrhea, previous exposure to suspected viral cases and healthcare workers. Independent predictors of mortality in NRS included age >60 years, diabetes, COPD, CKD, and decreased risk associated with previous exposure to suspected viral cases (**Supplementary Material**). After follow-up, 530 patients were still under domiciliary surveillance (6.2%), 127 remained hospitalized (1.5%), 146 were discharged after recovery (1.7%), 4 had voluntary discharge and 7,626 had completed follow-up with domiciliary surveillance without developing RS (89.2%).

### Profiling initially asymptomatic cases with SARS-CoV-2 infection

Amongst initially asymptomatic patients with SARS-CoV-2 infection (n=6,112), 185 were eventually hospitalized (3.0%), 12 required MVS (0.2%) and 29 required ICU admission (0.5%). We recorded 32 deaths (0.5%) amongst whom 11 deaths were ambulatory (0.2%) and 21 were hospitalized (0.3%). Predictors for severe COVID-19 included age >60 years, CKD, and reduced risk associated to previous exposure to suspected viral cases; predictors of lethality included age >60 years and CKD. As of August 19^th^ 2020, 415 patients were still under domiciliary surveillance (6.8%), 91 remained hospitalized (1.5%), 71 were discharged after recovery (1.2%), 2 had a voluntary discharge, and 5,501 had completed follow-up without developing symptoms which required hospitalization (90.0%).

### Predictors and outcomes for RS, NRS and asymptomatic cases

Compared to initially asymptomatic cases, symptomatic cases were associated with older age, reduced previous viral exposure, healthcare workers, diabetes, obesity, hypertension, CKD, COPD, asthma and smoking. Similarly, compared to NRS, RS cases were associated with asthma, immunosuppression, COPD, older age, male sex, diabetes, obesity, hypertension, reduced previous viral exposure with a reduced likelihood associated with smoking (**Figure 2**). Compared to initially asymptomatic cases, risk of severe COVID-19 was significantly higher for both NRS (HR 1.56, 95%CI 1.14-2.13) and RS cases (HR 4.78, 95%CI 3.67-6.21), adjusted for age, sex and comorbidities. Similarly, NRS and RS had higher mortality compared to asymptomatic cases (Breslow-Cox p<0.001, **Figure 2**); adjusting for age, sex and comorbidities, NRS (HR 2.32, 95%CI 1.57-3.43) or RS cases (HR 6.50, 95%CI 4.59-9.21) retained higher mortality compared to initially asymptomatic cases.

**Figure 2.**
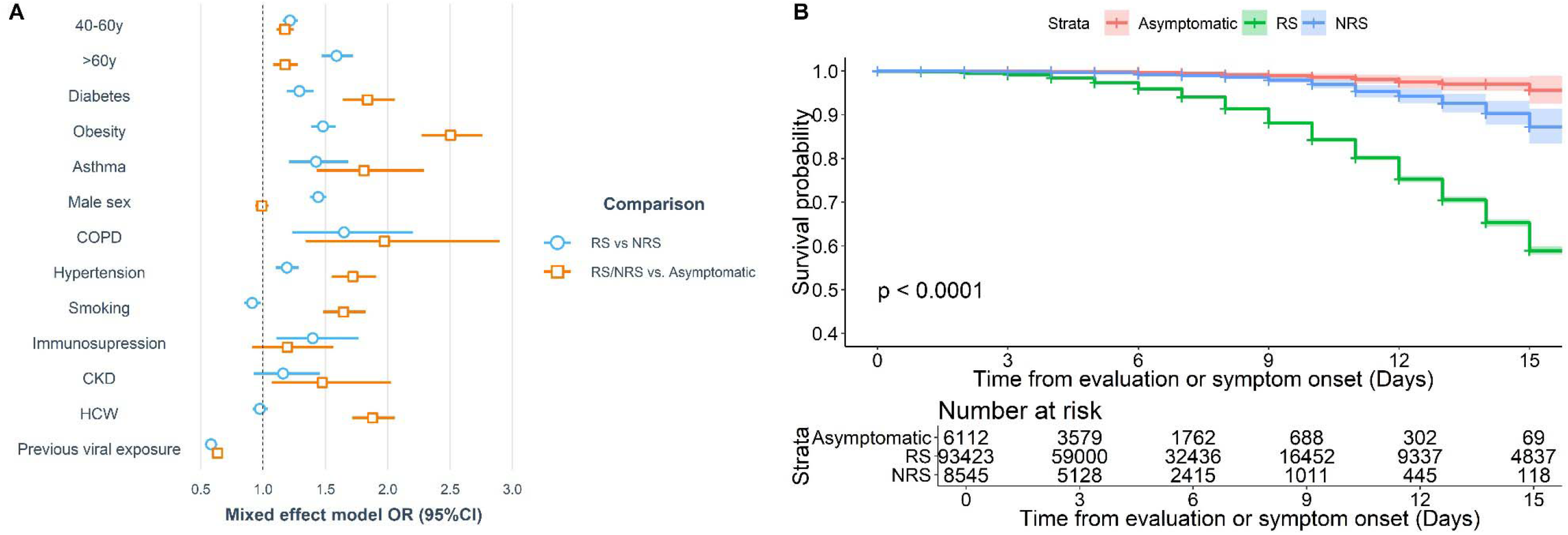
Factors associated with presence of RS compared to NRS, and RS/NRS compared to asymptomatic cases with SARS229 CoV-2 infection (A). We also show the and comparison of 30-day mortality rates between RS cases, NRS and asymptomatic SARS CoV-2 infections in Mexico City (B). <u>*Abbreviations*:</u> RS: Respiratory symptoms; NRS: Non-respiratory Symptoms; COPD: Chronic obstructive pulmonary disease; CKD: Chronic kidney disease; HCW: Health-care workers; Previous viral exposure: Prior contact with a patient with suspected viral infection.

## DISCUSSION

Here, we present a comprehensive report of COVID-19 cases with NRS and initially asymptomatic SARS-CoV-2 infections in Mexico City. Early detection of SARS-CoV-2 infections in initially asymptomatic patients or who present with NRS would help to reduce the spread of SARS-CoV-2 in Mexico, as these patients have lower risk of subsequent complications, as shown in this study, and would benefit the most from domiciliary treatment and prompt isolation. We also report a low prevalence of cases who were initially classified as asymptomatic but eventually were hospitalized or had outcomes related to severe COVID-19^8^. This data confirms observations from other populations regarding the course of COVID-19 cases with NRS and initially asymptomatic SARS-CoV-2 infections^3^. Our study also highlights the relevance of extending testing to cases with NRS and initially asymptomatic cases within the testing framework in Mexico; identifying asymptomatic cases is of importance given conflicting evidence on viral shedding for these cases, and data suggesting evidence of subclinical lung abnormalities which must be assessed longitudinally to rule out long-term impacts of SARS-CoV-2 infection ^9^. Notably, a consistent predictor of outcomes for NRS cases and initially asymptomatic cases were previous exposure to suspected viral cases, highlighting the relevance of contact tracing and prompt case identification but also confirming that cases presented in our study are not representative of all asymptomatic infections but mostly of those identified by contact tracing, likely underestimating rates of true asymptomatic cases.

Studies assessing natural history of initially asymptomatic SARS-CoV-2 infections highlighted the role of age and comorbidity in the subsequent development of symptoms, which is consistent with our observations^10^. Domiciliary surveillance in Mexico City focused on outcomes but not individual symptom onset, which might limit our ability to adequately characterize pre-symptomatic cases, particularly those who only develop mild symptoms and do not require hospital admission. Regardless, only a small fraction of initially asymptomatic cases developed severe COVID-19 after initial assessment, most of whom had underlying chronic comorbidities or were susceptible for COVID-19 complications attributable to unmasked cardiometabolic comorbidities. The eventual presentation of severe COVID-19 in initially asymptomatic cases might be attributable to a combination of decreased early immune response to SARS-CoV-2 infection and late enhanced pro-inflammatory responses in subjects with comorbidities ^9,11^. This might also extend to subjects with NRS with COVID-19 whom have previously been reported to delay seeking medical attention and have prolonged viral shedding compared with subjects with RS only ^12^.

Here, we assessed a thorough constellation of symptoms in one of the largest studies of NRS and initially asymptomatic SARS-CoV-2 infections. Additional limitations include nonassessment of atypical SARS-CoV-2 symptoms which might occur amongst otherwise asymptomatic cases, particularly in older adults, and the lack of systematized sampling, which does not allow for an accurate estimation for the rates of asymptomatic cases in Mexico City. Our results highlight the need to systematize definitions of asymptomatic cases and extend testing by contact tracing to detect asymptomatic SARS-CoV-2 infections as one of the mitigations strategies to reduce transmission of SARS-CoV-2 in Mexico City and Mexico in general.

## Data Availability

All data sources and R code are available for reproducibility of results at https://github.com/oyaxbell/covid_asymptomatic_cdmx.

## ACKNOWLEDGMENTS

NEAV, JPBL, and AVV are enrolled at the PECEM program of the Faculty of Medicine at UNAM. JPBL and AVV are supported by CONACyT. The authors would like to acknowledge the invaluable work of all of Mexico’s healthcare community in managing the COVID-19 epidemic. Its participation in the COVID-19 surveillance program has made this work a reality, we are thankful for your effort.

## AUTHOR CONTRIBUTIONS

Research idea and study design OYBC, NEAV, AVV, JPBL, CAFM, AMS; data acquisition: OYBC; data analysis/interpretation: OYBC, JPBL, NEAV, AVV, CAFM, AMS; statistical analysis: OYBC; manuscript drafting: OYBC, NEAV, AVV, JPBL, CAFM, AMS; supervision or mentorship: OYBC. Each author contributed important intellectual content during manuscript drafting or revision and accepts accountability for the overall work by ensuring that questions about the accuracy or integrity of any portion of the work are appropriately investigated and resolved.

